# Adult Patient Perspectives on Psychiatric Polygenic Risk Scores

**DOI:** 10.64898/2026.09.24.26363942

**Authors:** Craig W. McFarland, Amanda R. Merner, Page M. Trotter, Abigail C. Martinez, Lauren A. Ginn, Jonathan Frumovitz, Daphne M. Ayton, Takahiro Soda, Eric A. Storch, Stacey Pereira, Gabriel Lázaro-Muñoz

## Abstract

Polygenic risk scores (PRS) are rapidly developing, enabling estimates of genetic risk for developing psychiatric disorders. Psychiatric PRS are also increasingly entering consumer and clinical settings. Yet, little is known about how adults with psychiatric disorders perceive their potential use. Here, we conducted semi-structured interviews with 29 adults with psychiatric disorders to examine their perspectives on hypothetically receiving psychiatric PRS for themselves and their current or future children. Key themes emerged around perceived utility, motivations and deterrents to genetic testing, interpretation of PRS, and anticipated responses to results. In particular, participants expressed hope that PRS could support self-understanding, inform better psychiatric care, and enable earlier recognition of difficulties for their children. Participants however raised multiple concerns, including those of privacy, stigma, discrimination, eugenics, and the risk of self-fulling prophecies. Together, these findings offer critical empirical evidence to inform guidance on the responsible use of psychiatric PRS in clinical and consumer settings.

---

Psychiatric disorders are among the leading causes of disability worldwide, with rates of diagnosis and treatment alike increasing across populations globally (Arias et al., 2022; GBD 2019 Mental Disorders Collaborators, 2022; Vigo et al., 2016). Amid escalating public health concern, there has been substantial investment in early identification, risk prediction, and personalized treatment strategies within psychiatry (Meehan et al., 2022; Sun et al., 2025).

Genetic risk prediction in particular is rapidly advancing in psychiatry. Expanding genomic datasets, together with advances in artificial intelligence and machine learning, have enabled the development of models to estimate individualized risk (Meehan et al., 2022; Sun et al., 2025). Among these tools, polygenic risk scores (PRS) have emerged as a focus in psychiatric research. PRS quantify genetic liability by summing the effects of many common variants associated with a condition, offering a probabilistic estimate of risk compared to the general population (Chatterjee, Shi, & García-Closas, 2016; Lewis & Vassos, 2020).

PRS have now been developed for numerous major psychiatric conditions, including attention-deficit hyperactivity disorder (ADHD), bipolar disorder, major depressive disorder, opioid-use disorder, and schizophrenia, based on findings from large-scale genome-wide association studies (Deak et al., 2022; Demontis et al., 2023; Howard et al., 2019; Lam et al., 2019; Stahl et al., 2019; Trubetskoy et al., 2022). Early studies suggest that these scores can identify individuals at elevated risk. For instance, those in the highest percentile of depression PRS have been found to carry a two-fold increased risk of depression, rising to about 30% compared to 15% in the general population (Murray et al., 2021), and up to a 32-fold increased risk of recurrence and psychiatric comorbidity (Als et al., 2023). As psychiatric disorders are early in their onset and widespread in their prevalence, there is strong motivation to explore tools like PRS that could be used to support early detection and intervention (GBD 2019 Mental Disorders Collaborators, 2022; Kessler et al., 2007; Solmi et al., 2022). To these ends, clinicians have noted that PRS could be used in the future to identify individuals at heightened genetic risk and inform individualized prevention, detection and treatment strategies (Merner et al., 2024).

Psychiatric PRS are also becoming more accessible through commercial genetic testing services and emerging clinical platforms. Direct-to-consumer genetic testing has grown rapidly, with the global market projected to expand more than sevenfold from $8.84 billion in 2023 to $64.7 billion by 2033 (Phillips et al., 2018; Vision Research Reports, 2024). Concurrently, psychiatric PRS are becoming increasingly accurate, driven by the expansion of datasets and methodological advances (Meehan et al., 2022; Sun et al., 2025). As a result, psychiatric PRS may become an increasingly accessible and widespread instrument in both clinical and commercial settings.

Despite advances, uncertainties remain concerning how psychiatric PRS are perceived and should be applied in real-world settings. Much of the existing literature has centered on the technical development and validation of PRS, with comparatively less attention paid to their real-world regulation, reception, and use. Recently, a small but growing body of research has begun to explore perspectives and concerns among clinicians toward PRS, including child and adolescent psychiatrists (Ginn et al., 2026; Merner et al., 2024; Pereira et al., 2022; Trotter et al., 2025). Yet, no study has systematically examined how adult psychiatric patients themselves would perceive and anticipate responding to PRS information in hypothetical scenarios.

Thus, there is limited empirical insight into how these individuals evaluate the relevance of psychiatric PRS, anticipate their potential utility, or consider the kinds of concerns, uncertainties, and implications that such scores could entail. In addition, psychiatric PRS currently have modest predictive utility and limited clinical validity (Torkamani et al., 2018; Wray et al., 2021). Without clear patient input on how they might perceive, interpret, and act upon PRS information, applying PRS in psychiatry risks miscommunication, misplaced expectations, and unintended harms. Such concerns are especially salient in psychiatry, where diagnostic categories are heterogeneous, symptom trajectories variable, and treatment response uncertain (Feczko et al., 2019).

Concerns about how patients interpret and respond to psychiatric PRS are further heightened by the persistent stigma associated with mental illness and the documented effects of genetic framing. Historical eugenic misuse of “heritability” demonstrates the risks of reductionist interpretations (Lombardo, 2018). Genetic explanations of psychopathology have been shown to increase perceptions of patients as abnormal, less capable of recovery, diminished in agency, and even less than fully human (Lebowitz & Ahn, 2014; Phelan et al., 2006). Individuals with psychiatric symptoms and known genetic risk are also perceived as more dangerous and impaired, and studies suggest that they are subject to greater social distance, including reduced willingness to engage in close relationships such as marriage (Jorm & Oh, 2009; Jorm et al., 2012; Kvaale et al., 2013; Phelan, 2005). These dynamics are both socially and clinically significant, as stigma can interfere with self-identification of symptoms, help-seeking, treatment engagement, recovery, and self-perception (Corrigan, 2004; Henderson et al., 2013; Clement et al., 2015). Finally, concerns also extend to the potential for discrimination based on psychiatric genetic risk, underscoring the importance of legal, ethical, and policy safeguards (Brannan et al., 2019). Understanding how patients themselves make sense of psychiatric PRS is therefore essential for maximizing benefits and anticipating and mitigating potential harms as these tools move closer to clinical and consumer implementation.

Prior research also illustrates how concerns about genetic risk extend into considerations regarding current and potential children. Individuals with a family history of mental illness often weigh the possibility of transmitting psychiatric conditions when making decisions about having children. For example, approximately one-third of respondents in prior studies reported reduced willingness to have genetically-related children out of concern of inheriting bipolar disorder (Meiser et al., 2007; Trippitelli et al., 1998). Similar worries about inherited vulnerability have also been observed among mothers in treatment for substance use disorders, over half of whom expressed concerns about their children’s risk for addiction (Keller et al., 2024). Some adults expressed a desire to learn whether they or their offspring may carry genetic markers linked to psychiatric disorders, viewing this knowledge as important for guiding family decisions (Austin et al., 2006). Most recently, these considerations of genetic risk have been examined in the context of psychiatric PRS, with approximately half of adults with psychiatric conditions expressing interest in using the emerging tool for family planning and embryo screening (Ginn et al., 2026). Understanding how patients interpret psychiatric PRS may therefore yield potential insights into broader personal and familial considerations involved in psychiatric genetic testing.

To address this gap, this study investigates how adults with a range of psychiatric conditions may interpret, respond to, and act upon polygenic risk scores for mental health conditions. This study makes three primary contributions. First, it offers the first empirical analysis of how adults with psychiatric disorders perceive the potential use of psychiatric PRS, including their views on hypothetically receiving this information for themselves and for their current or future children. Second, it identifies a set of comprehensive themes and subthemes that help characterize how patients make sense of and may engage with polygenic risk information if they were to receive this testing. Third, it provides actionable insights to support the development of communication strategies and implementation approaches that reflect patient experiences and priorities. Together, these contributions address a critical gap in psychiatry and can inform guidelines that promote the responsible integration of psychiatric PRS into clinical and consumer contexts.

## Methods

### Approach

We interviewed adult patients receiving outpatient psychiatric care (*n* = 30) to examine their perspectives on psychiatric PRS. Participants were eligible if they were at least 18 years old and were currently receiving treatment for a psychiatric condition. We included both patients with children and those intending to have children to capture perspectives on the potential use of PRS for both current and future children. Participants were recruited through online advertisements or direct patient outreach at approved clinical sites. This study was approved by the Baylor College of Medicine Institutional Review Board (protocol number H-50235), and all participants provided informed consent.

We developed a semi-structured interview guide based on previous research on psychiatric genetics, PRS, and stakeholder perspectives on the implications of using genetic information in psychiatric care (Merner et al., 2024; Pereira et al., 2022; Trotter et al., 2025). To support informed responses, participants were given a plain-language explanation of PRS, including how these scores are derived and how they might be used. See Supplementary Materials for the complete semi-structured interview guide.

### Data Cleaning and Analysis

Interviews were audio-recorded, transcribed, and de-identified to ensure anonymity prior to analysis. We used a hybrid deductive and inductive approach to analyze the data (Fereday & Muir-Cohrane, 2006). Researchers trained in qualitative analysis (ARM, PMT, LG, ACM, JF) first deductively identified core topics based on our primary research questions (Crabtree & Miller, 1992). Primary coders then independently applied these thematic codes to the interview transcripts. Secondary coders then reviewed the coded material and collaborated with primary coders to resolve any discrepancies through consensus discussions.

Following consensus coding, excerpts within each thematic code were exported for further analysis. We analyzed participant responses to all interview questions related to perspectives on the utility of psychiatric PRS, perceived benefits and harms, interest in testing, and anticipated behavioral responses. Responses to questions focused specifically on reproductive decision-making were excluded from analysis, as those are addressed in a separate manuscript (Ginn et al., 2026).

At this stage, a primary coder generated summaries of each excerpt using an *in vivo* approach that emphasized participants’ own language and framing (Bernard, 2018). These summaries were then synthesized into broader interpretive themes that captured higher-order patterns across responses (Lindgren et al., 2020). Where applicable, subthemes were identified to represent more specific or nuanced viewpoints within each overarching theme. A secondary coder independently reviewed the thematic abstractions for coherence and accuracy, and any disagreements were resolved collaboratively to ensure consistency (Creswell, 2018). Finally, a separate researcher reviewed frequencies of theme and subtheme occurrence to assess their salience and distribution across participants. Within each theme, subthemes are generally presented in order of their frequency in the Results section. Thematic saturation was reached (Bernard, 2018). Importantly, frequency counts are included to promote transparency, providing readers with information to discern how many participants raised a given theme instead of relying on imprecise terms like “some” or “many.” This approach enables readers to gauge the relative prominence or salience of themes across different perspectives (e.g., whether only one participant raised a specific issue or if the concern was shared across the sample). These frequency counts, however, are not intended to imply generalizability.

## Results

We interviewed 30 adults currently receiving psychiatric treatment over the age of 18. One interview was excluded from data analysis after determining that they were not eligible due to not currently receiving psychiatric care, leaving a total of 29 patient interviews that were included in the final analysis. Participants were predominantly female (n = 19; 65.5%) and White (n = 21; 72.4%). The detailed demographics of the final analytic sample are presented in Table 1.

**Table 1.** Participant Demographics.

| Participant Characteristics | (n = 29) |
| --- | --- |
| **Gender** |  |
| Female: | 19 (65.5%) |
| Male: | 10 (34.5%) |
| **Race** |  |
| White/European American: | 21 (72.4%) |
| Asian or East Asian: | 2 (6.9%) |
| Black/African American: | 4 (13.8%) |
| More than one race: | 1 (3.4%) |
| Other * | 1 (3.4%) |
| American Indian/Alaska Native: | 0 (0.0%) |
| **Ethnicity** |  |
| Hispanic/Latino/a/e: | 20 (69.0%) |
| Not Hispanic/Latino/a/e: | 9 (31.0%) |
| **Age** |  |
| Average (SD) | 30.84 (SD=9.50) |
| Range | 19.0 to 54.7 |
| **Education** |  |
| Grade 12 | 6 (20.7%) |
| Trade School | 1 (3.4%) |
| Associate's degree | 1 (3.4%) |
| Bachelor's degree | 14 (48.3%) |
| Master's degree | 5 (17.2%) |
| Doctoral degree | 2 (6.9%) |
| **Income** |  |
| Less than \$25,000 | 4 (13.8%) |
| \$25,000 to \$34,999 | 1 (3.4%) |
| \$35,000 to \$49,999 | 2 (6.9%) |
| \$50,000 to \$74,999 | 2 (6.9%) |
| \$75,000 to \$99,999 | 3 (10.3%) |
| \$100,000 to \$149,999 | 3 (10.3%) |
| \$150,000 or more | 12 (41.4%) |
| Not reported | 2 (6.9%) |
| **Child status** |  |
| Has children | 11 (37.9%) |
| Does not yet have children, but plans to | 18 (62.1%) |
| **Diagnosis †** |  |
| OCD | 14 (48.3%) |
| Anxiety | 14 (48.3%) |
| Depression | 7 (24.1%) |
| ADHD | 4 (13.8%) |
| PTSD | 2 (6.9%) |
| Bipolar Disorder | 2 (6.9%) |
| Misophonia | 1 (3.4%) |
| Trichotillomania | 1 (3.4%) |
| **Years since Diagnosis** |  |
| 1 to 5 years | 5 (17.2%) |
| 6 to 10 years | 7 (24.1%) |
| 11 to 15 years | 4 (13.8%) |
| 16 to 20 years | 2 (6.9%) |
| 21 to 30 years | 6 (20.7%) |
| Greater than 30 years | 5 (17.2%) |

\* The participant who noted "Other" in Race category identified as "Latina"; this is also reflected in the "Hispanic/Latino/a/e" category above.
† Frequencies add up to greater than total sample (n = 29) due to the presence of comorbid conditions in 12 participants.
NOTE: Table 1 originally appears in Ginn et al. (2026), which reports on a different subset of interview data from the same sample. Reuse is intended to support consistency and transparency in reporting.

### Perceived Utility of Psychiatric PRS

When asked about whether and how psychiatric PRS could be useful, nearly all patients viewed it as a tool that could enhance how they parent, plan, and care for their child’s wellbeing.

The most commonly cited benefit was that PRS could help inform or improve medication and treatment decisions (18/29). For instance, several (12/29) envisioned PRS as a tool for early intervention, particularly in contrast to their own past experiences with delayed or crisis-driven care. One patient emphasized this concern, stating, “*I just feel like instead of waiting until everything came to crisis and a hospitalization, diagnosis, treatment, care, maybe things could have been better for her. We could have completely avoided a crisis”* (AP15). Similarly, some patients (5/29) anticipated that the information may be useful for medical staff treating the child, or even preventing the development of the condition altogether (2/29). Many patients (13/29) also believed PRS could help them prepare proactively for the possibility of psychiatric conditions in their child. Several (10/29) highlighted PRS as potentially helping to narrow down diagnoses, providing clarity where there may have been multiple overlapping presentations.

PRS was also framed as a tool for better parenting overall. Patients noted that PRS may improve their parenting strategies (8/29), help them to identify and track symptoms (8/29), and potentially prevent severe episodes or crises through early awareness (6/29). One patient captured this proactive mindset, remarking, “*…this is the beginning of all of this, but I would still get tested anyway. And I would still have my kid get tested anyway. Because it’s something new. At minimum, it’s like a tarot card reading. At maximum, it will help me influence my child’s life*” (AP8). Further, some parents suggested that having this information could help normalize psychiatric risk within the family (5/29) and even prevent stigma (5/29), enabling candid conversations about mental health and illness. As one explained, *“So I know and I can raise my kids saying, ‘Hey, this is what’s going on… you have a disposition towards [a psychiatric condition], but we can manage it and it’s normal.’ …it would be normalizing it”* (AP1).

Others anticipated value in sharing PRS information with schools or care teams (5/29), such as to help attain proper learning accommodations, or to support ongoing management at home (5/29). While less frequently mentioned, participants also imagined benefits such as gaining more information about their child’s behavior and personality (3/29), reducing self-guilt or blame (1/29), and increasing education surrounding psychiatric conditions (1/29).

Finally, some patients suggested that PRS could serve as a tool for self-understanding (4/29), or for helping them explain themselves to others and legitimize their experience (1/29): “*I think that the benefit of the testing, one, for responding to people, knowing how to talk to them. And the second one would be for proof, like to say, ‘Look, they have this.’”* (AP23).

### Perceived Disutility of Psychiatric PRS

While many participants recognized potential benefits of psychiatric PRS, there was a robust expression of concern about their risks and ethical implications.

There was near-unanimous concern among participants (26/29) that psychiatric PRS might lead to discrimination or being treated differently based on results. Two participants specifically invoked concerns of eugenics, with one noting that “*…in general, it could definitely have issues with parents who then start to treat their child more poorly because of it, or stuff along the lines of eugenics*” (AP29). Many (12/29) also voiced concerns that PRS results could negatively impact self-image or self-esteem, or even worsen their conditions by reinforcing negative expectations (e.g., self-fulfilling prophecy; 5/29). As one patient stated, *“…any disease that you could have is just a small part of who you really are. So for me, is that my kids would think that, that’s all they are, which is totally not true”* (AP17).

Others expressed discomfort with the idea of children knowing too much about their psychiatric risk: *“[The child] not knowing is also not bad. If anything, it could be detrimental because I do believe that the more you think about something, the more you manifest it… ignorance is bliss.”* (AP13). Moreover, some feared that knowing their PRS may negatively impact their parenting (5/29) or their family unit at large (3/29). One patient articulated this worry about labels becoming limiting: *“…it runs the risk of pigeonholing them in a certain diagnosis. Diagnoses can be helpful and I know that likelihood is not a diagnosis. Right? But even boxing someone within a specific label, I think, creates a lot of harm…”* (AP14).

Another anticipated concern was data privacy (21/29), with parents voicing uncertainty about who would access the results, how securely data would be stored, and how academic institutions, companies or insurers might use the information. Nearly half the participants (14/29) opposed including the hypothetical PRS data in academic records, citing potential misuse or misunderstanding in educational settings. Nearly a fourth (7/29) feared that acquiring PRS data may negatively impact their medical treatment and insurance, with some (4/29) explicitly opposing inclusion of PRS in their medical records. Others noted concerns of legal ramifications (2/29) and commercialization (2/29).

A minority of participants explicitly rejected some of the aforementioned concerns, stating they were not personally worried about discrimination (4/29), privacy (2/29), or harm to parenting (2/29). Lastly, over a third (10/29) questioned the meaningfulness of the results altogether, noting that PRS merely reflects risk, not diagnosis.

### Motivations for Psychiatric PRS Testing

#### a. Motivations for Psychiatric PRS Testing for Children

When asked hypothetically whether they would want to know the psychiatric PRS for their current or future child, participants expressed a range of motivations for wanting this information.

The most common theme related to motivation was a desire to use PRS as an educational tool to learn about psychiatric conditions that might affect their child (21/29), with many participants emphasizing wanting to be as informed as possible. Some patients (7/29) expressed interest in using PRS for early detection or identification of psychiatric symptoms in their children, as well as early treatment and intervention (9/29), and the majority indicated interest in using PRS information to support lifestyle modifications to potentially prevent the condition. One patient expressed this desire clearly: “*If I knew that they had a certain psychiatric condition, I would be able to either get them in touch with doctors sooner or talk to their teachers or whatever I could do as a parent to make their life easier*” (AP20). Many (14/29) felt it was their parental responsibility to request testing if it would be helpful to their child and some (4/29) also hoped PRS could support parental preparation or advocacy, such as helping them better care for and speak up for their child in healthcare or school settings. Other notable parental motivations included wanting to create an environment at home that was open to discussing mental health (14/29) and helping parents simply understand their child better (8/29). Additionally, nearly half of the patients (14/29) mentioned a known family history of psychiatric disorders as a likely motivator for acquiring PRS.

#### b. Motivations for Psychiatric PRS Testing for Self

A majority of participants (20/29) also expressed interest in hypothetically undergoing PRS testing for themselves. These reflections were often framed in terms of better preparation: almost a third of patients (9/29) felt that PRS might be desirable in helping to prepare for future psychiatric events. One participant described the regret of missing that opportunity earlier in life: “*Had I known… I would’ve had a chance to know, to learn about the symptoms of it and then be aware of it… I could’ve caught it 20 years ago, instead of after having it for 20 years*” (AP26). Select patients (4/29) voiced potential value in using PRS to positively reframe their conditions, such as understanding them as biologically based rather than due to personal failings. Others (7/29) envisioned sheer curiosity as a driver. A few also noted the relevance of personal family history (3/29), and some identified interest in specific disorders like schizophrenia (2/29) and bipolar disorder (2/29).

### Deterrents for Psychiatric PRS Testing

When asked hypothetically whether they would want to know their child’s psychiatric PRS, participants also described reasons that would deter them from pursuing such information.

#### a. Deterrents for Testing in Self

The most common deterrent to hypothetically pursuing testing for oneself was that psychiatric PRS might increase emotional burden. Several participants (6/29) anticipated that receiving genetic risk results could cause anxiety or lead them to become adversely hypervigilant. One patient explained, *“…if for some reason I find out I am 60% likely to be depressed or experience depression, I don’t want that statistic to be looming over me. Rather, I just want to lead my life as best as I can and keep, I guess, leading my little lie that I’m perfectly normal…*” (AP13). Others (5/29) worried that PRS might offer unnecessary or unhelpful information, particularly if they already had a formal diagnosis or felt adequately informed about their condition.

Some participants (4/29) envisioned the likelihood of a self-fulfilling prophecy as a potential deterrent, in which high-risk results might reinforce existing negative beliefs or shape expectations in unhelpful ways. “*I would just be so obsessed with whether or not I was showing symptoms of it that it would maybe even accelerate me getting it,*” one patient admitted. “*But if I just didn’t know, then I could just be blissfully ignorant*” (AP14). Others felt testing might not be beneficial because they were already confident in their understanding of their mental illness (4/29). Smaller groups of patients described a general preference to address problems as they emerge (3/29) and skepticism toward genetic determinism (2/29).

#### b. Deterrents for Testing in Children

When asked about hypothetical interest in obtaining psychiatric PRS testing for their children (current or future), some participants expressed ambivalence or reservations. A few (3/29) directly voiced ambivalence, while others (3/29) suggested they would be hesitant as PRS results might be harmful if misunderstood or overemphasized. Several participants (5/29) worried that knowing a child’s genetic risk might negatively affect parenting, such as leading to altered expectations or more anxious decision-making.

Some feared that PRS test results could shape how the child views themselves. Two patients anticipated that risk information might inadvertently influence a child’s identity development or behavior by creating a self-fulfilling prophecy. Others voiced hesitations about increasing the child’s own anxiety or sense of burden (2/29). A few participants (3/29) expressed a lack of interest in pursuing the hypothetical testing for specific conditions or in contexts where they felt the information would not be actionable, while others (4/29) were skeptical about the accuracy of PRS testing.

### Interpretation of Psychiatric PRS Results

Participants demonstrated a wide range of interpretations when responding to how they would make sense of psychiatric PRS results.

The most common interpretation was that a low PRS does not rule out the possibility of developing a psychiatric condition (19/29). In alignment with the background information provided, many participants reiterated that these scores reflect probabilities, not certainties, with one noting that “*…it’s not a guarantee. It’s just a risk*” (AP11). Others (11/29) viewed a low PRS as indicating a lower likelihood of developing a condition, while remaining aware that it was not definitive.

Some participants (9/29) emphasized the importance of environmental factors, including family context, trauma, and life stressors, in interpreting a PRS. One patient in particular noted that for disorders that are “*very much moreso genetically predisposed, I want to know about that more*” (AP22). A small number (2/29) interpreted a low PRS as a positive indicator that their child was doing well. Only one participant (1/29) expressed the belief that a low PRS meant their child would not develop a psychiatric condition.

One participant voiced concern that a low score could lead to complacency, potentially discouraging attention to early symptoms or mental health needs. No participants described a high or low PRS as inherently harmful, or a high PRS as more helpful.

### Actions in Response to Psychiatric PRS Results

When asked how they would respond hypothetically to psychiatric PRS results for their children, participants described actions ranging from no action to seeking outside resources.

The most commonly anticipated action was to share the results with a doctor (11/29), with participants emphasizing the importance of clinical interpretation and professional guidance. Many (10/29) also described a driving desire to be prepared for what might come, including emotionally bracing themselves, having a plan in place, or preparing family members. Others indicated they might begin monitoring for specific symptoms (9/29), and several (8/29) anticipated using the information to support early intervention or pursue educational resources related to psychiatric conditions. One patient explained this mindset as a layered approach to readiness: “*[Psychiatric PRS] would potentially allow us to push our child into resources and services early to identify factors or to just provide support just in case. And also to educate them as well”* (AP8).

Participants also anticipated seeking support (7/29), adopting preventive measures (7/29), and making lifestyle or environmental changes (7/29). Some said they would focus treatment based on results (7/29) or share PRS information with schools or educators (5/29). For some, this involved strategic communication: “*I would want to see if that could be used to get an expedited 504 or whatever… I might white lie to the school. ‘We have this in our family, so could you look out for these things if you start to notice…’ I want to make sure we could have early intervention…”* (AP10). Others, however, were more cautious: “*I wouldn’t share it with the teachers unless it was a diagnosis, and even then I’m 50-50. We live in a small town and they bored. They just love to chit chat*” (AP15).

Others suggested that they might use psychiatric PRS to advocate for their child (5/29), learn coping strategies (5/29), or regarded any information as helpful irrespective of risk level (5/29). Several participants imagined using PRS to guide parenting decisions (3/29), start therapy (3/29), or inform reproductive decisions (3/29).

Disclosure to children, however, was a point of divergence: some participants (4/29) explicitly said they would not anticipate sharing results with their child, while others (2/29) expressed a possible intention to disclose. Others expressed conditional approaches, noting that disclosure might depend on symptoms (3/29) or perceived preferences of the child (1/29). One patient clarifies, *“Again, it goes back to telling the kids if they need to know… But I’m not going to just go throw it around like the Friday newspaper”* (AP22).

Lastly, some indicated that they might keep the results private (5/29), while a few said they would hypothetically share them with immediate family (3/29) or spouses/co-parents (2/29). Others noted that any action they might take would be contingent on factors such as the specific condition (1/29), the PRS score itself (1/29), or the presence of symptoms (1/29). Only one participant (1/29) indicated they would hypothetically take no action in response to PRS results.

## Discussion

This study is the first to qualitatively examine how adults who receive outpatient psychiatric care interpret and envision the use of psychiatric polygenic risk scores (PRS) for themselves and their children. Whereas prior research work has focused primarily on clinicians and technical development, there remains a critical gap in understanding how those with lived psychiatric experience may respond to this emerging technology. In this study, patients provided several novel perspectives on anticipated benefits, concerns, and informational needs related to psychiatric PRS. In addition, participants highlighted how they might interpret and use this information in parenting, decision-making, and personal reflection, while also raising concerns about stigma, psychological burden, and misuse of results.

Participants frequently described potential benefits of PRS in ways that extended beyond treatment decisions. Many envisioned PRS as a tool that could help them monitor psychiatric risk in their children, prepare for future care needs, or make sense of their own psychiatric experiences. These perceived benefits often involved emotional insight, proactive parenting, and long-term planning, suggesting that participants evaluated PRS through a broader personal and relational lens that went beyond just clinical utility. This aligns with prior findings that parents report high perceived value for PRS-based reports for their children (Terek et al., 2022).

These patient-centered findings contrast with prior research on child and adolescent psychiatrists, who have typically evaluated PRS through the lens of clinical applicability and emphasized the current lack of clinical utility as a primary barrier to implementation (Merner et al., 2024; Pereira et al., 2022; Trotter et al., 2025). Clinicians often prioritize clinically actionable outcomes when determining whether to adopt a new tool, whereas many patients expressed interest in PRS even in the absence of clear clinical interventions (Soda et al., 2021). Clinicians’ skepticism is supported, provided the modest effect sizes and low individual-level predictive power of current psychiatric PRS (Ni et al., 2021), and a recent review finds that the clinical utility of PRS in the near future may be limited to narrow clinical questions (Smith et al., 2023). Even so, studies show that clinicians may nevertheless order PRS when requested by patients or parents, despite their own doubts about utility (Merner et al., 2024; Pereira et al., 2022; Soda et al., 2023).

By contrast, patients expressed interest in psychiatric PRS for personal reasons, describing ways that genomic information could meaningfully contribute to emotional preparedness, behavioral planning, or personal understanding (Mackley et al., 2017). These different interpretations reflect broader questions in the field regarding the nature of utility and whose definition should guide implementation (Owens et al., 2023). Together, these findings reinforce the perspective to address utility as a multidimensional concept that incorporates both medical and personal considerations (Torkamani et al., 2018; Smith et al., 2021).

Participants also imagined concerns about how PRS results could be misinterpreted or misused. Participants were particularly worried about stigma and discrimination from family members, schools, or employers. These concerns are well-supported by prior research. For example, adolescents with mental health conditions often report stigmatizing experiences from family, peers, and school personnel (Moses, 2010), while adults may encounter similar stigma in the workplace (Follmer & Jones, 2018). One study found that imagining receiving a low-percentile polygenic score for educational attainment led participants to rate both themselves and a hypothetical classmate lower in self-esteem, competence, and educational potential, suggesting risks of stigma and self-fulfilling prophecies (Matthews et al., 2021). Genetic information may further intensify these risks, with evidence suggesting that such disclosures can strain familial and social relationships (Horstkötter et al., 2012), lower parental expectations for a child’s future (Corcoran et al., 2005), or negatively affect a child’s developing identity and sense of potential (Rosenthal & Jacobson, 1968; Acheson & Papadima, 2023).

Several participants also feared that receiving a positive result could increase anxiety or contribute to the onset of psychiatric symptoms. This concern comports with prior studies indicating that genetic feedback can influence psychological wellbeing and increase stress (Dar-Nimrod et al., 2013; Dinç & Terzioglu, 2006; Wilhelm et al., 2009), and that receiving PRS results can elicit negative emotional reactions (Peck et al., 2022). Prior research also demonstrates that individuals often grapple with whether and how to disclose genetic testing results, the potential for discrimination, and the impact of genetic information on self-perception and family dynamics (Klitzman, 2012).

Many emphasized a preference for receiving results in a clinical context if they were to seek PRS, citing concerns about accuracy, data privacy, and the importance of having a trusted professional to help interpret the information. Prior work has shown that patients prefer to receive genetic results in person, especially when those results may raise questions or emotional reactions (Ashtiani et al., 2014; Stuttgen et al., 2020). Similar challenges have been noted in pediatric contexts, with one study finding that parents experienced confusion in interpreting PRS-based reports without clinician guidance (Terek et al., 2022). This need for informed result disclosure places added responsibility on clinicians, particularly psychiatrists, to deliver and explain PRS effectively.

Yet, prior literature suggests limited clinician preparedness in psychiatry. A recent study found that fewer than half of surveyed experts in psychiatric genetics were able to correctly answer all objective knowledge questions about PRS (Moorthy et al., 2023), highlighting the lack of training and potential for misinterpretation even among specialists (Klitzman et al., 2014; Pereira et al., 2022). Additionally, a survey of psychiatrists and neurologists found that nearly half believed genetic testing could cause psychological harm, and many viewed their training as insufficient to support patients effectively (Salm et al., 2013). This gap in clinician knowledge, paired with patients’ reliance on clinician guidance for managing psychiatric disorders, underscores the need for expanded training as PRS become more widely available in psychiatric contexts. Accordingly, clinicians themselves have identified education as a key step to ensuring the responsible integration of PRS (Merner et al., 2024).

Compared with their stronger motivation to seek PRS testing for their children, participants were more ambivalent about hypothetically testing for themselves. Some for instance envisioned being curious about their genetic risk or illness origins, while others anticipated being reluctant due to concerns about determinism, emotional burden, or self-image. The envisioned concerns are consistent with empirical findings that exposure to genetic explanations can reduce individuals’ confidence in their ability to cope with illness (Lebowitz & Ahn, 2018) and lower their perceived potential in academic or social domains (Matthews et al., 2021). In one study, informing older adults that they carried a heightened genetic risk for Alzheimer’s disease led them to judge their memory more negatively and perform worse on objective memory tests compared to those with the same risk who were unaware of it (Lineweaver et al., 2014).

Anticipated responses to PRS results varied widely. Although most understood PRS as probabilistic, many worried that other stakeholders, including educators or insurers, might treat the information as definitive. Prior research supports this concern, showing that even among professionals, PRS can be misunderstood or overinterpreted (Donohue et al., 2021). A smaller group of participants noted that low-risk results could also be meaningful, providing reassurance or indicating that further preventive efforts were not necessary. This aligns with Stuttgen et al. (2020), who found that virtually all participants in their study reported negative genomic screening results as valuable. Yet, research also shows that individuals who endorse biogenetic explanations for conditions such as schizophrenia and depression often display lower social acceptance of affected persons (Schomerus et al., 2013), suggesting that PRS disclosure could unintentionally reinforce stigma.

Several limitations must be considered when interpreting these findings. First, the study is subject to sample biases. Participants were predominantly White, female, college-educated, and actively receiving psychiatric care. These characteristics may reflect a population that is more informed about, or more open to, discussing psychiatric genetics than the broader population of adults with psychiatric conditions. Second, the study may also reflect response bias, as individuals who chose to participate may have had a particular interest in genetic testing or mental health advocacy, which could influence the perspectives discerned. These biases are particularly salient as there are well-documented disparities in how individuals from different racial and ethnic backgrounds perceive and interact with mental health care. For instance, stigma surrounding psychiatric disorders is also more commonly reported in racial and ethnic minority communities (Eylem et al., 2020). Research also demonstrates racial and ethnic disparities in access to and use of mental health care, with Black, Hispanic, and Asian individuals less likely than White individuals to receive professional care (Manuel, 2018; Cook et al., 2017; McGuire & Miranda, 2008). Similar patterns have also been observed among children (Rodgers et al., 2022). These differences likely shape both access to care and attitudes toward psychiatric risk, and they should be addressed in future studies using more diverse and representative samples. Finally, participants responded to hypothetical scenarios rather than reflecting on personal experiences with psychiatric PRS results. While this approach supports exploration of perceptions and reasoning, it may not accurately capture how individuals would behave or respond in real-world contexts.

Despite these limitations, this study offers critical insight into how adults with psychiatric disorders may interpret, respond to, and act upon PRS for mental health conditions. As such, it provides a foundation for future research and can inform the development of implementation strategies, educational resources, and clinical practices that are responsive to patient priorities and concerns. Future studies should examine how individuals respond to PRS information in actual clinical or consumer settings and assess downstream behavioral, psychological, and relational outcomes. Additionally, our findings frequently centered on how PRS might be used in relation to participants’ children, underscoring the importance of understanding how youth themselves perceive and respond to this information. Research that explores adolescent perspectives directly is essential, particularly given the potential for PRS to influence identity development, academic expectations, and social dynamics during a critical developmental stage.

Psychiatric PRS are complex and evolving tools. Their effects will depend not merely on statistical accuracy and predictive power, but critically on how individuals with lived experience of psychiatric conditions interpret, respond to, and integrate this information into their lives. As these novel technologies move closer to clinical and consumer settings, centering patient perspectives will be essential for anticipating how these tools will function outside the research setting, and for guiding their ethical and effective implementation in psychiatric care.

## Funding

The research reported in this article was supported in part by the Dana Foundation grant for the Neurotech Justice Accelerator at MGB (NJAM), a Dana Center for Neuroscience & Society, as well as by the National Institutes of Health (R01MH128676). The views expressed in this article are those of the authors and not necessarily those of the funders.

## Disclosures

GLM is founder of Fidelia, a services firm that works with health technology companies. EAS reports receiving research funding to his institution from the International OCD Foundation, Wellcome Trust, and NIH. EAS receives direct funding from the International OCD Foundation as well as MHNTI for providing trainings on treating obsessive-compulsive disorder with psychotherapy. EAS co-founded Rethinking Behavioral Health which provides training and consultation in the treatment of obsessive-compulsive disorder and related conditions. EAS was a consultant for Brainsway and Biohaven Pharmaceuticals in the past 36 months. EAS owns stock options less than $5000 in NView (for distribution of the Y-BOCS and CY-BOCS) and receives royalties from OCD Scales LLC (for distribution of the Y-BOCS and CY-BOCS). EAS receives book royalties from Elsevier, Wiley, Oxford, American Psychological Association, Guildford, Springer, Routledge, and Jessica Kingsley. TS reports receiving funding from the NIH to his institution for work not related to this manuscript. In the past three years, TS has received travel support from the Florida Association for Behavior Analysis and The Minnesota Northlands Association for Behavior Analysis to discuss the importance of cross-disciplinary clinical collaboration.

## Ethics approval

This study was approved by the Baylor College of Medicine Institutional Review Board (protocol number H-50235), and all participants provided informed consent.

## Data availability statement

The de-identified aggregate data supporting the findings of this study are available from the corresponding author upon reasonable request. The survey instrument is provided as Supplementary Material.

## Notes

### Author Declarations

IRB of Baylor College of Medicine gave ethical approval for this work.

