## Supplemental Table 1 for "Adult Patient Perspectives on Psychiatric Polygenic Risk Scores"

Adult Psychiatric Patient Interview Guide

[verbal consent]

***Rapport Building and Background***

Before we begin the main portion of the interview, we’d like to ask you a few questions about your experience with your treatment so far.

1. What is your current psychiatric diagnosis?
   a. When did you first begin experiencing symptoms?
   b. When were you diagnosed?
2. Tell me about your family.
   a. Do you have any children?
      i. How many?
      ii. How old are they?
      iii. (If not addressed) Are they your biological children?
   b. Tell me about your parents.
      i. Are you aware of any family members who also have [participant’s condition]?
      ii. Are you aware of any other psychiatric diagnoses any of your family members have?
3. Do you think a person’s genes contribute to how likely the person is to develop a psychiatric condition?
   a. How much?
4. Just to remind you, we don’t expect you to have any previous knowledge about genetics. Have you heard about genetic testing for risk of psychiatric conditions before?

For this next part of the interview, we’re interested in hearing your thoughts about a new type of genetic testing, called polygenic risk scores. “Poly” means “many”, so the term “polygenic” just means that there are many genes involved in a particular condition. Conditions like psychiatric disorders involve many genes, so they are often called “polygenic conditions.” There is now a way to look at many genes that are involved with psychiatric disorders to create a “score” for the chances a person might have a particular condition now or in the future. We’ll specifically be talking about using these polygenic risk scores to look at risk for psychiatric disorders—for example, depression, obsessive-compulsive disorder (OCD), and schizophrenia. As I mentioned before, we don’t expect you to be an expert or have any knowledge about genetics. And I also want to stress that you [and your child] won’t be actually getting this testing done. We’re just interested in hearing your thoughts and learning about your perspectives on these topics.

***PRS for Self***

5. Have you ever had any experiences with genetic testing before? Please tell me about it.
   a. (If they don’t mention in response to question above)  Has your doctor ever mentioned or suggested ordering genetic testing?
   b. (If yes to question above) Can you tell me more about that?
6. Do you think you would want to know your genetic risk for psychiatric conditions? Why/Why not?

***PRS for Child***

Note: If participants reported not having children, questions in this section were prefaced with “If you had children”.

7. (If they say child[ren] do not have psychiatric conditions or symptoms in background section) How likely do you think it is that your child will develop a psychiatric disorder?
8. Do you think you would want to know your child’s polygenic risk score for psychiatric conditions? Why/Why not?
   a. Which specific psychiatric conditions do you think you would want to know about?
   b. Are there some psychiatric conditions you would NOT want to know about?
9. How do you think this information could be useful?
   a. Do you think it could be beneficial for your child’s treatment? In what ways?
10. How do you think this information could be harmful to your child, if at all?
    a. Some people have raised concerns about the potential for this information to be used to discriminate against someone. What do you think about this?
11. What information do you think you would need to help you decide whether to receive your child’s [children’s] polygenic risk scores for psychiatric conditions?
    a. (Alternative phrasing) If a doctor were to offer this type of testing for your child, what information would you want to know before you decided whether or not to get the test done?
12. If you were to get a test like this for your child done and it showed they were at a higher risk of a psychiatric condition….
    a. What do you think that would mean [for your child’s risk of developing the condition]?
    b. What do you think you would do with this information?
    c. Would you share this information with your child?
    d. How would you tell them about the results?
    e. When would you tell them about the results?
    f. How do you think knowing this information might impact them?
    g. (If not addressed) Do you think it could have an emotional impact on them?
    h. How do you think knowing this information might impact you?
    i. (If not addressed) Do you think it could have an emotional impact on you?
13. If your child were to get a test like this done and their results showed your child was NOT at a higher risk of a psychiatric condition…
    a. What do you think that would mean [for your child’s risk of developing the condition]?
    b. What do you think you would do with this information?
    c. Would you share this information with your child?
    d. How would you tell them about the results?
    e. When would you tell them about the results?
    f. How do you think knowing this information might impact them?
    g. (If not addressed) Do you think it could have an emotional impact on them?
    h. How do you think knowing this information might impact you?
    i. (If not addressed) Do you think it could have an emotional impact on you?
14. Do you think knowing your child had a high PRS for a psychiatric condition would change anything about your parenting style?
    a. For instance, some people are worried that this type of information could cause parents to decrease their expectations for their child. What do you think about this?
    b. What impact, if any, might this information have on your family as a whole?
15. Who would you share your child’s PRS results with, if anyone? Why?
    a. Would it depend on what the results were?
16. Is there anyone you would not want to have access to your child’s PRS results? Why?
17. How would you feel about PRS results being part of your child’s medical record?
18. How would you feel about the use of your child’s PRS results outside the medical setting (e.g., employers)?
19. How appropriate do you think it would be for schools to be able to access PRS results?
20. How would you feel about PRS results being part of your child’s school records?
21. Are you familiar with any laws in the US that protect against genetic discrimination?
22. So far, we’ve discussed your perspectives surrounding ordering these tests from your child’s doctor. How would you feel if you were able to order this type of testing directly from a company like 23-and-me? Would you want to do that? Why?/Why not?
    a. (If they would be interested in direct-to-consumer testing) If you were to order this type of testing, what do you think you would do with the test results?
23. So far, we’ve talked about PRS testing for psychiatric conditions. Do you think you would want to know your child’s polygenic risk score for other, non-psychiatric conditions, like heart disease or diabetes? Why/Why not?
24. What information do you think you would need to help you decide whether to receive your child’s polygenic risk scores for non-psychiatric conditions?
    a. (Alternative phrasing) If a doctor were to offer this type of testing for your child for non-psychiatric conditions, what information would you want to know before you decided whether or not to get the test done?
25. Would you be interested in generating polygenic scores for educational attainment for your child? Why?/Why not?

***Genetics and PRS Impact on Reproductive Decisions***

26. Some doctors believe individuals knowing their genetic risk for a psychiatric condition may influence their reproductive decision-making. Do you think knowing you had a high PRS would influence your decision to have children?
27. Do you think knowing one of your children had a high PRS might influence your decision about having other children?
28. Do you think that having a psychiatric condition in any way influenced your decision about whether or when to have children?
29. It is possible to do genetic tests on embryos created through *in vitro* fertilization, or IVF. Soon, it may be possible to test these embryos to learn each embryo’s genetic risk of developing a psychiatric condition. Potential parents could use this information to choose which embryo to implant into the mother when using IVF. What do you think about using this information to select embryos for implantation?
30. If you were planning on having [more] children and you could use this technology, do you think you would want to know about your embryo’s risk for a psychiatric condition?
    a. (If yes) What do you think you would do with this information?

***Concluding remarks***

31. Are there any other thoughts you have on these topics that we didn’t cover yet that you’d like to share?
